# Improved visualization and segmentation of the choroid plexus using double inversion recovery MRI

**DOI:** 10.64898/2026.08.13.26360360

**Authors:** Markus Lauerer, Julian McGinnis, Cornelius Berberich, Tun Wiltgen, Einar August Høgestøl, Pål Berg-Hansen, the MultipleMS consortium, Jan S Kirschke, Bernhard Hemmer, Mark Mühlau

## Abstract

**Background:** Choroid plexus (CP) volume is an emerging magnetic resonance imaging (MRI) biomarker in various disorders of the central nervous system (CNS). However, clinical translation is hindered by methodological heterogeneity and inconsistent anatomical coverage. Double inversion recovery (DIR) – a sequence providing dual-tissue suppression – is a promising candidate to improve CP segmentation.

**Methods:** The dataset included 93 scans across healthy subjects and individuals with multiple sclerosis (MS), divided into a training set (*n* = 63), an internal test set (*n* = 20), and an external test set (*n* = 10). First, relative CP signal intensity and tissue contrast ratios on DIR were compared against fluid-attenuated inversion recovery (FLAIR) and T1-weighted (T1w) sequences (pre- and post-contrast). Reproducibility of manual CP segmentations was assessed via intraclass correlation coefficients (ICCs). Subsequently, we developed a 3D nnU-Net model for CP segmentation based on manually labeled DIR masks. Model performance was evaluated against manual segmentation using spatial overlap and volumetric error metrics. Finally, we compared our DIR-based model against three publicly available T1w- or FLAIR-based tools by assessing slice-wise volume distributions and voxel-wise density maps.

**Results:** DIR demonstrated the highest CP signal intensity and most consistent tissue contrast among evaluated MRI sequences (*p* < 0.001). Intra- and inter-rater agreement for manual CP segmentations was robust (ICC = 0.92 and 0.83, respectively). The trained nnU-Net achieved high internal accuracy (Dice = 0.82) independent of scanner, diagnosis, or absolute CP volume, and generalized well to the external test set (Dice = 0.75). Compared to public T1w- and FLAIR-based models, DIR-based approaches (nnU-Net and manual) yielded significantly larger CP volumes (*p* < 0.01). Axial volume distribution analysis attributed this difference to a distinct bimodal profile in DIR segmentations, more fully capturing the CP inside the temporal horn of the lateral ventricle (*p* < 0.001 against T1w- and FLAIR-based models).

**Conclusions:** By leveraging the superior tissue contrast of DIR, our nnU-Net model achieves highly accurate CP segmentation that generalizes across scanners and captures the inferior extent of the C-shaped structure often missed by conventional models. This may improve standardization of CP volumetry and allow for more reliable studies in CNS disorders.

## Background

The choroid plexus (CP) is a richly vascularized brain structure lining the cerebral ventricles. Previous studies suggest its involvement in various aspects of brain development and homeostasis, including the production of cerebrospinal fluid (CSF), metabolite clearance, secretion of neurotrophic factors, and neurogenesis [1, 2]. Furthermore, the monolayer of epithelial cells delimiting the CP forms the blood–CSF barrier, an interface that plays a central role in regulating immune cell trafficking in the central nervous system (CNS). This functional diversity has prompted investigations into its potential involvement in various CNS pathologies, ranging from inflammatory to neurodegenerative disorders [3].

In contrast to molecular and cell biology studies, macroscopic examinations of the CP using magnetic resonance imaging (MRI) are a relatively recent development. Here, increased CP volume has emerged as a potential surrogate marker for pathological processes in a wide range of CNS disorders, including multiple sclerosis (MS), dementia, Parkinson’s disease, amyotrophic lateral sclerosis, stroke, depression, and schizophrenia [4]. The significance of this observation remains unclear, however, not least because the correlations between CP volume and disease progression, as well as clinical scores, differ considerably across studies. This inconsistency may be at least partially attributed to the variety of approaches to CP segmentation. Apart from the various techniques used – spanning manual, semi-automated, and deep learning-based solutions – there is a notable lack of consensus regarding the preferred MRI sequence; studies currently utilize T1-weighted (T1w), T2-weighted, fluid-attenuated inversion recovery (FLAIR), contrast-enhanced (CE) T1w sequences, or combinations thereof [5].

This methodological heterogeneity is reflected in the high variability of CP volumes found in the literature. A recent large-scale study of more than 5500 subjects documented mean adult CP volumes ranging from 1126 mm^3^ (females 19–30 years) to 1921 mm^3^ (males 81–90 years) [6]. In contrast, other studies of healthy adults have reported mean values lower than 800 mm^3^ [7–9] or exceeding 2800 mm^3^ [10–12], suggesting that methodological choices significantly influence results beyond age-related changes. Moreover, 3D reconstructions indicate that current CP segmentations often fail to capture the inferior extent of its C-shaped structure as it curves from the ventricular body through the atrium and into the temporal horn, where the plexus narrows and becomes closely bounded by surrounding brain parenchyma [13–16].

Double inversion recovery (DIR) is an MRI pulse sequence designed to simultaneously suppress signals from two distinct tissue types [17]. It is primarily utilized in MS imaging to improve the visualization of cortical lesions [18, 19], but has also proven useful for identifying white matter (WM) lesions [20, 21]. Although DIR has not yet been evaluated for CP segmentation, its contrast properties make it an ideal candidate for this task. The unique microstructure of the CP (high vascularity, protein-and lipid-rich matrix) allows it to retain a hyperintense MRI signal on DIR, while the surrounding tissues that typically complicate both manual and automated delineations are effectively nulled (CSF, WM) or significantly attenuated (gray matter, GM).

In this study, we investigated signal properties and contrast behavior of the CP on DIR compared to conventional MRI sequences. We further evaluated the internal and external performance of a deep learning model trained specifically for this task and assessed its anatomical CP coverage in relation to other publicly available segmentation models.

## Methods

### Data acquisition and image postprocessing

The study comprised three distinct datasets with a total of 93 individual MRI scans (Table 1; Figure 1 provides a study flowchart): an internal training set (*n* = 63), an internal test set (*n* = 20), and an external test set (*n* = 10).

**Figure 1:**
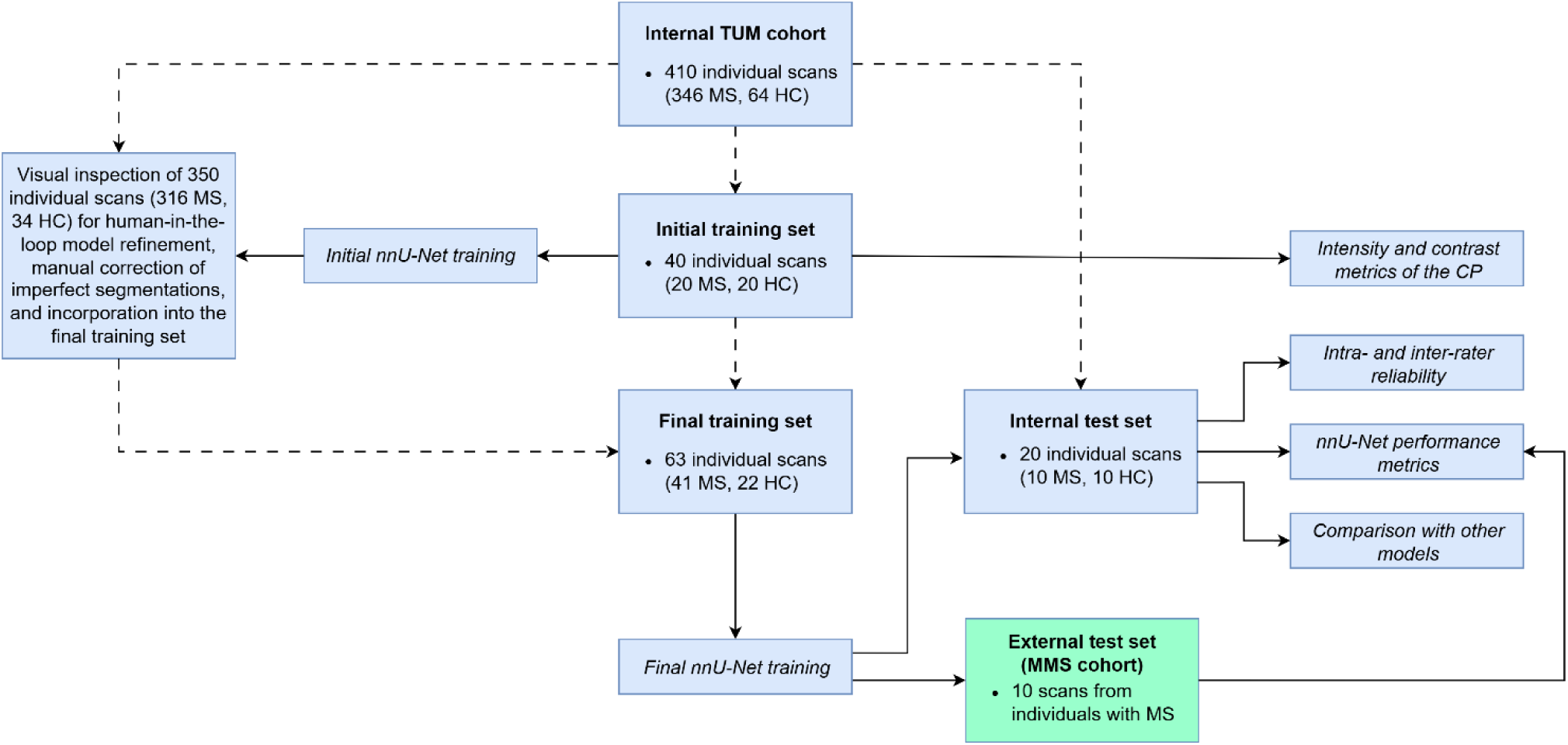
Study flowchart. Dashed lines represent connections between datasets; solid lines represent applications of datasets to different analyses. CP, choroid plexus; HC, healthy controls; MMS, Multiple MS; MS, multiple sclerosis; TUM, Technical University of Munich.

**Table 1:**
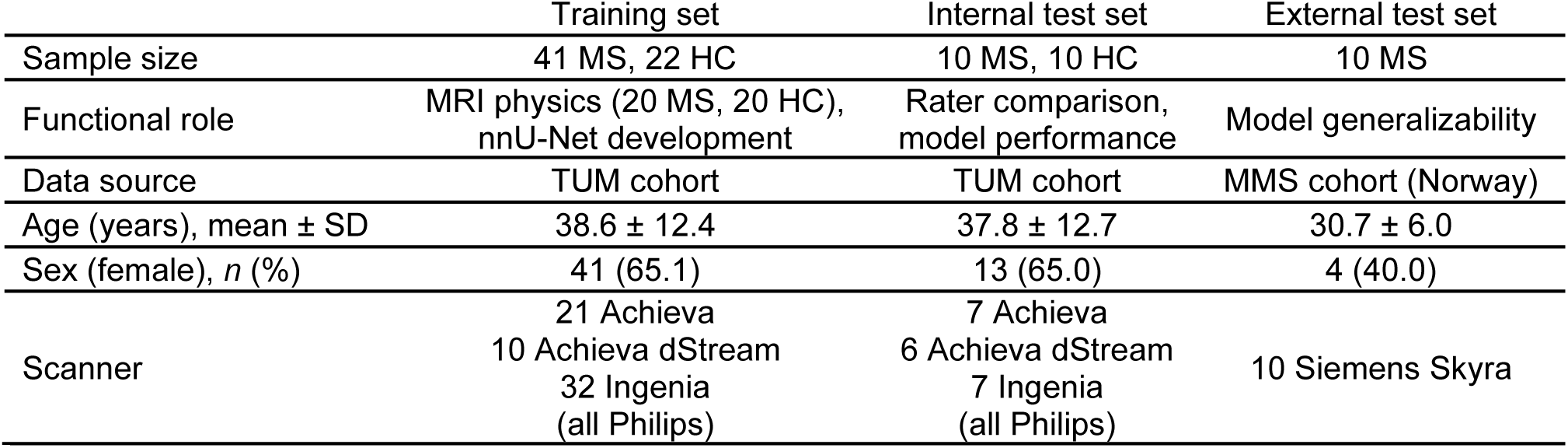
Characteristics of the study datasets. HC, healthy controls; MMS, Multiple MS; MRI, magnetic resonance imaging; MS, multiple sclerosis; SD, standard deviation; TUM, Technical University of Munich.

|  | Training set | Internal test set | External test set |
| --- | --- | --- | --- |
| Sample size | 41 MS, 22 HC | 10 MS, 10 HC | 10 MS |
| Functional role | MRI physics (20 MS, 20 HC),<br>nnU-Net development | Rater comparison,<br>model performance | Model generalizability |
| Data source | TUM cohort | TUM cohort | MMS cohort (Norway) |
| Age (years), mean ± SD | 38.6 ± 12.4 | 37.8 ± 12.7 | 30.7 ± 6.0 |
| Sex (female), $n$ (%) | 41 (65.1) | 13 (65.0) | 4 (40.0) |
| Scanner | 21 Achieva<br>10 Achieva dStream<br>32 Ingenia<br>(all Philips) | 7 Achieva<br>6 Achieva dStream<br>7 Ingenia<br>(all Philips) | 10 Siemens Skyra |

Internal dataset scans were acquired at the Technical University of Munich (TUM) University Hospital. For healthy controls (HC), scans were obtained either during clinical routine or previous research studies, whereas scans for participants with MS were collected as part of an ongoing cohort study.

Additionally, an extended pool of 350 scans (316 MS, 34 HC) from TUM University Hospital was used to iteratively improve segmentation accuracy via a human-in-the-loop approach, with 23 of these scans (21 MS, 2 HC) ultimately incorporated into the final training set. Subjects with MS were intentionally included to control for potential interference of MS lesions with CP segmentation, as both structures display hyperintense signals on DIR sequences. Since age- and MS-related brain atrophy and subsequent ventricular enlargement may influence CP volume metrics, we deliberately selected a heterogeneous cohort across the adult lifespan.

Internal MRI data (TUM) were acquired across three scanners between 2013 and 2023 (Achieva, Achieva dStream, and Ingenia; Philips Healthcare, Best, NL) to capture a realistic degree of hardware and protocol heterogeneity. Imaging protocols comprised 3D DIR, pre- and post-contrast 3D T1-w magnetization-prepared rapid gradient echo (MPRAGE), and 3D FLAIR sequences (see Supplementary table 1 for sequence parameters). All four sequences were rigidly co-registered using Greedy [22] with the T1w scan as reference, followed by T1w-based skull-stripping of each co-registered sequence using HD-BET [23]. This study component was approved by the TUM Ethics Committee (reference number: 5848/13), and participants provided written informed consent to use their data for research purposes.

The external test set was obtained on a Siemens Skyra scanner between 2018 and 2020 from a single center (Norway) of the prospective Multiple MS study cohort [24]. MRI parameters are detailed in Supplementary table 1.

### Tissue signal intensity and contrast metrics

Manual segmentations for CP masks and extractions of signal intensity values were performed in 3D Slicer [25]. In accordance with previous literature, segmentations were restricted to the CP inside the lateral ventricles, as CP tissue within the third and fourth ventricles is comparatively small and difficult to reliably delineate on MRI. Whole brain, GM, WM, and CSF masks were acquired using the Sequence Adaptive Multimodal SEGmentation (SAMSEG) tool with simultaneous lesion labeling to avoid potential tissue misclassification [26]. MS lesion masks were generated using LST-AI [27].

All tissue masks were morphologically eroded by one voxel using a 3D cross-shaped structuring element (six direct orthogonal neighbors) to avoid measurement bias from partial volume effects. This process isolated a conservative tissue core that is likely to be located within the tissue of interest across all co-registered sequences (DIR, T1w, CE T1w, and FLAIR). For anatomically narrow segments, a conditional safeguard was implemented to preserve original voxels if erosion would otherwise reduce the local tissue thickness to zero – a concern particularly relevant to the irregular structure of the CP.

Z-score normalized mean intensity values were calculated according to the following formula: *(SI_CP_ – SI_brain_) / SD_brain_*. Here, *SI_CP_* represents the mean signal intensity of the CP, while *SI_brain_* and *SD_brain_* denote the mean signal intensity of the whole brain and its standard deviation (SD), respectively.

The contrast ratio between the CP and adjacent tissues was defined as *|SI_CP_ – SI_T_| / (SI_CP_ + SI_T_)*, where *SI_CP_* is the mean signal intensity of the CP and *SI_T_* is the mean signal intensity of the contrasted tissue. It ranges between 0 (no contrast between tissues) and 1 (maximum contrast between tissues) [20]. To assess the consistency of contrast ratios across tissue types, coefficients of variation (CoVs) were calculated as *SD_CR_ / mean_CR_*.

### Intra- and inter-rater reliability

For all 20 scans from the internal test set, the CP of the lateral ventricles was manually segmented in the coronal plane of the 3D DIR sequence by an experienced neurologist (M.L.; see Figure 2 for a representative segmentation). The scans were then re-segmented by the primary rater after a 10-week interval (to avoid familiarity bias) and independently segmented by an experienced neuroradiologist (C.B.), respectively.

**Figure 2:**
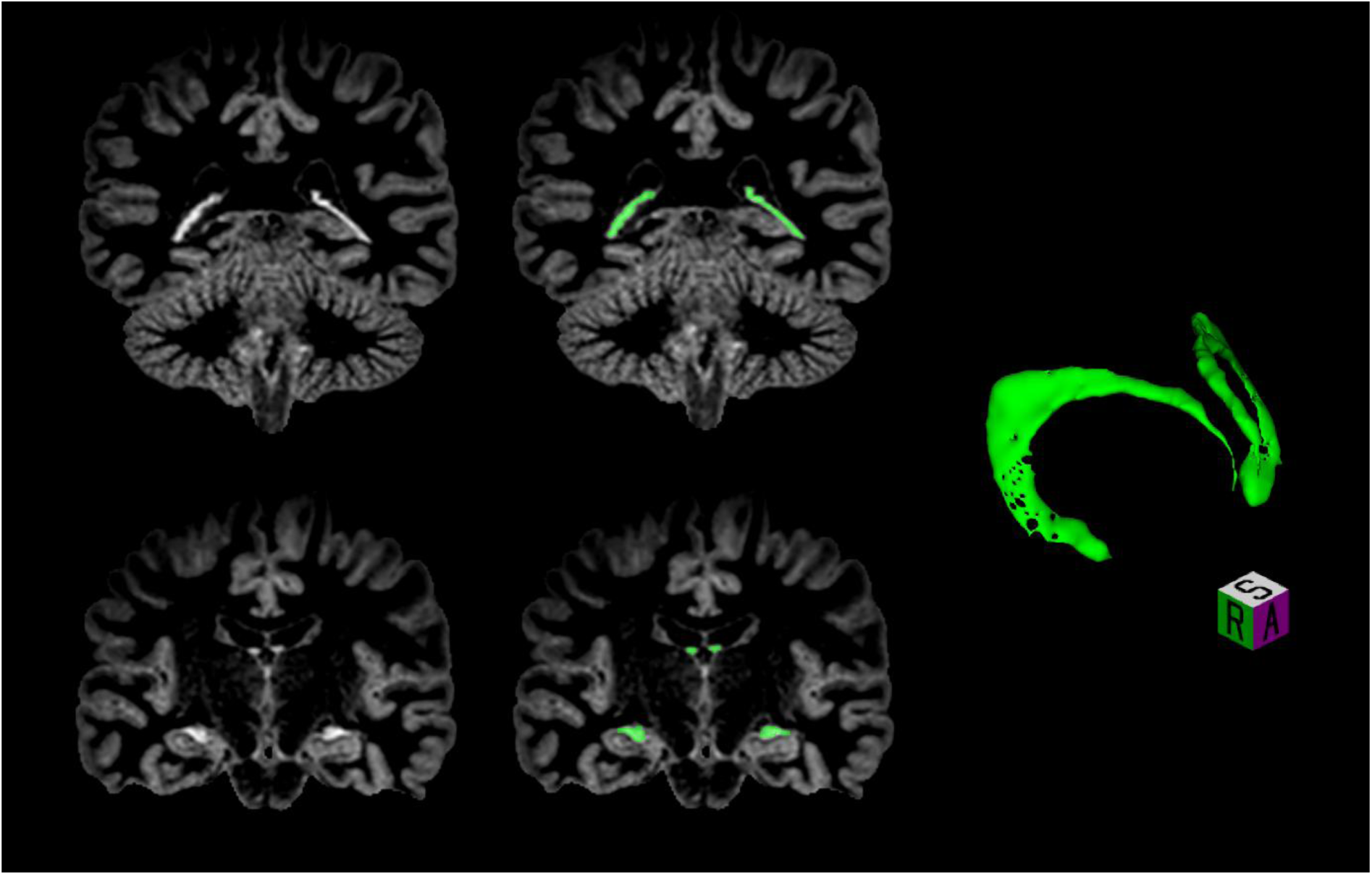
Representative manual CP segmentation on DIR. Coronal slices showcase the distinct anatomical subregions of the choroid plexus across the ventricular body, atrium, and temporal horn of the lateral ventricles (left, manual segmentations overlaid in green). The reconstructed 3D surface model (right) illustrates the C-shaped geometry of the segmented structure. A, anterior; CP, choroid plexus; DIR, double inversion recovery; R, right; S, superior.

Intraclass correlation coefficients (ICCs) based on a single-measure, two-way random-effects model with absolute volume agreement were calculated to assess both intra- and inter-rater reliability – ICC(2,1) according to the nomenclature by [28]. Additionally, the Dice similarity coefficient (DSC), mean surface distance (MSD), Hausdorff distance (HD), and absolute relative volume difference (RVD) were determined in each case.

### Deep learning model training and performance evaluation

Automated segmentation of the CP was performed using the nnU-Net framework, a self-configuring architecture widely utilized in biomedical image segmentation [29]. To maximize model robustness and capture anatomical edge cases, we employed an iterative, human-in-the-loop labeling strategy for training data curation. An initial base model was trained on 40 DIR scans manually segmented by M.L. This preliminary model was subsequently deployed for inference on a larger dataset of 350 unannotated scans. Following visual inspection of the resulting segmentations, 23 scans with imperfect labeling of the CP were identified (see Supplementary figure 1 for examples). These 23 scans were manually corrected by M.L. and integrated back into the primary training pool, resulting in a final training cohort of 63 individual scans.

Model training and internal validation were executed using the default 3D full-resolution configuration of the nnU-Net framework. The requirement for a standalone, held-out validation cohort was purposefully bypassed in favor of nnU-Net’s built-in 5-fold cross-validation mechanism. During this procedure, the 63 scans from the training dataset were iteratively partitioned; for each fold, four partitions were utilized for weight updating, while the fifth served as an internal validation set for hyperparameter optimization and prevention of overfitting. The final segmentation model was generated as an ensemble of the five trained cross-validation folds to maximize spatial prediction stability.

To evaluate model performance, the nnU-Net was applied to the 20 scans of the internal test set. The following metrics were then determined in relation to manual CP segmentations (done by M.L.): DSC, recall, precision, MSD, HD, RVD, and absolute RVD. Additionally, model generalizability was assessed by applying the nnU-Net to an external DIR dataset of 10 MS patients (different country and MRI scanner) and determining the same performance metrics.

### Spatial distribution of CP segmentations by different models

To evaluate the anatomical CP coverage of our DIR-based approach in relation to other existing solutions, the manual annotations and nnU-Net outputs were compared against three publicly available CP segmentation tools: two T1w-based models by Visani et al., 2024 [30] and Li et al., 2025 [6], one FLAIR-based model by Eisma et al., 2024 [14]. Although Visani et al. recommend fine-tuning their model with manual, T1w-based segmentations prior to applying it to a new dataset, we chose to forego this step to evaluate model performance “out-of-the-box”. Two distinct spatial analysis pipelines were implemented to assess the precise anatomical localization of segmented volumes using the internal test set (*n* = 20).

<u>Slice-wise volume distribution:</u> To map the axial distribution of CP volume along the superior-inferior axis, all evaluated segmentation masks were resampled to the native grid of the manual segmentation using nearest-neighbor interpolation. The bounding box of the CP along the axial plane was defined individually for each subject based on the spatial extent of the manual segmentation, expanded by one slice superiorly and inferiorly to ensure boundary inclusion. Absolute slice positions along the z-axis were then converted into a normalized, continuous coordinate system ranging from 0% (most superior slice) to 100% (most inferior slice). For each normalized slice coordinate, the absolute cross-sectional volume (in mm³) was calculated.

<u>Voxel-wise MNI heatmaps:</u> To visualize the spatial density of CP segmentation across the test set, group-level heatmaps were generated in standard anatomical space. For each subject, the native T1-weighted structural scan was non-linearly registered to the MNI152 template (version 2009c) using Symmetric Normalization (SyN) implemented in the Advanced Normalization Tools Python suite (ANTsPy) [31]. The resulting forward deformation fields were subsequently applied to all binary segmentation masks using nearest-neighbor interpolation to preserve structural boundaries. The warped MNI-space masks were then aggregated across the entire test set; voxel-wise overlap maps were computed by dividing the cumulative sum of active voxels by the total number of subjects, yielding a continuous density spectrum (0.0 to 1.0).

### Statistical analyses and software

Comparisons of CP z-score signal intensities and contrast metrics were conducted using a repeated-measures analysis of variance (ANOVA). Post-hoc testing was performed using Dunnett’s test to specifically compare all other MRI sequences against the DIR reference.

A multivariable linear model (with Type III sum of squares) was constructed to test whether nnU-Net performance was dependent on scanner type, subject status (HC vs. MS), or CP volume (taken from manual segmentations). Additionally, potential proportional bias of the nnU-Net performance was evaluated using a Bland-Altman regression model, assessing the relationship between the segmentation volume differences and their averages.

Volumetric and spatial comparisons among the four automated segmentation models (nnU-Net + three publicly available models) and manual annotations were conducted using a within-subject, repeated-measures framework. CP volumes were compared via repeated-measures ANOVA, followed by Tukey’s Honestly Significant Difference (HSD) post-hoc tests for all pairwise comparisons. To investigate relative volumetric variance across the models, we employed a modified Levene’s test approach adapted for repeated measures: volumes were log-transformed, and absolute residuals from the model-specific medians were analyzed using a linear mixed-effects model (with the subject as a random intercept), followed by Tukey’s HSD test for pairwise comparisons.

Differences in the spatial distribution of the CP segmentations along the superior-inferior axis were quantified by calculating the area under the cumulative distribution function (CDF) for the slice-wise volumes of each model. This process of extracting the integral of a spatial mass curve across a normalized axis has its origins in oceanography and geology [32], but provides a robust, dimensionless metric to assess the relative volume distribution of any 3D structure. Values for the area under the CDF were compared across models using a repeated-measures ANOVA and post-hoc Tukey HSD testing.

Unless otherwise specified, continuous variables are reported as mean ± SD. Two-sided *p* values < 0.05 were considered statistically significant. Statistics and plotting were performed in *R* (version 4.5.3), using the *emmeans* (version 2.0.3), *ez* (version 4.5.0), *psych* (version 2.6.3), and *tidyverse* (version 2.0.0) packages. Image processing was performed using *Python* (version 3.13.12) and its packages *antspyx* (version 0.6.3), *nibabel* (version 5.4.2), and *numpy* (version 2.3.5).

## Results

### Sequence-specific signal behavior of the CP

DIR demonstrated the highest *z*-score normalized signal intensity of the CP among all evaluated MRI sequences (2.99 ± 0.34; all *p* < 0.001; Table 2). It also yielded the most pronounced CP-to-WM contrast (0.74 ± 0.05; all *p* < 0.001) as well as a CP-to-GM contrast (0.32 ± 0.04) superior to T1w and FLAIR (both *p* < 0.001) and equal to the one achieved by CE T1w imaging (*p* = 0.95). While the CP-to-CSF contrast was substantial on DIR (0.51 ± 0.12), it was significantly lower compared to all other sequences (all *p* < 0.001). Overall, DIR demonstrated the most consistent contrast behavior across these three main tissue types, exhibiting the lowest CoV (0.43 ± 0.06; all *p* < 0.001; Supplementary figure 2 shows example scans for each sequence). In the subcohort of 20 subjects with MS, CP-to-lesion contrast on DIR (0.15 ± 0.08) was not significantly different compared to CE T1w (*p* = 0.60) or FLAIR (*p* = 0.90), but was lower than the contrast observed on T1w scans (*p* = 0.001).

**Table 2:**
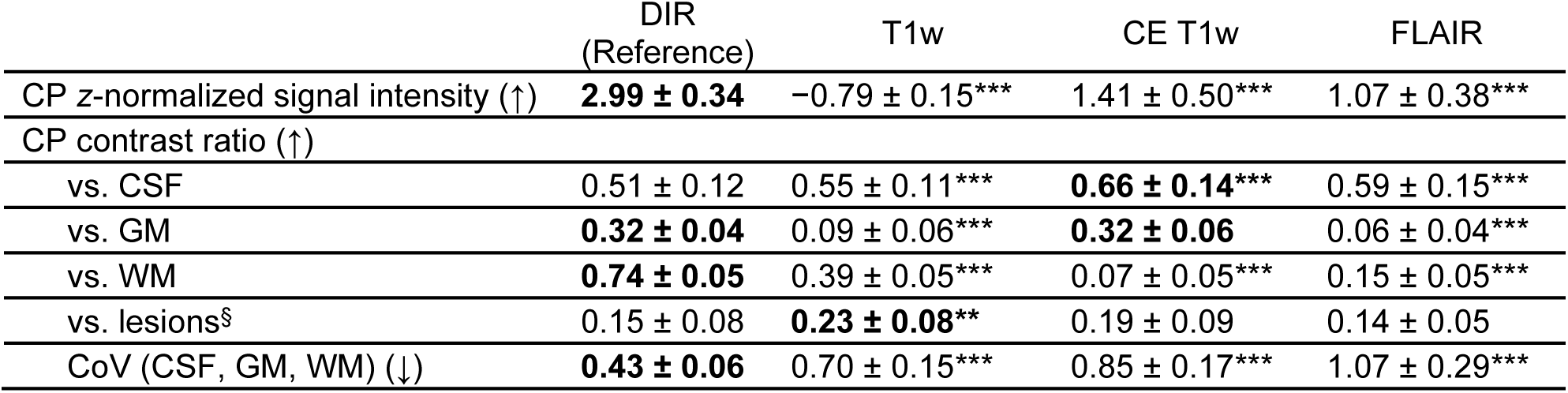
Signal and contrast metrics of the CP across different MRI sequences (*n* = 20 MS, 20 HC). Values are expressed as mean ± standard deviation. Arrows indicate the target direction for each variable. Bold values indicate the numerically optimal sequence within each row. Main effects were assessed via repeated measures analysis of variance (ANOVA), followed by post-hoc Dunnett’s tests using DIR as the designated reference group. ^§^ within MS subcohort (*n* = 20); ** *p* < 0.01 compared to DIR; *** *p* < 0.001 compared to DIR. CE, contrast-enhanced; CoV, coefficient of variation; CP, choroid plexus; CSF, cerebrospinal fluid; DIR, double inversion recovery; FLAIR, fluid-attenuated inversion recovery; GM, gray matter; HC, healthy controls; MRI, magnetic resonance imaging; MS, multiple sclerosis; T1w, T1-weighted; WM, white matter.

### Manual segmentation reliability and performance of the nnU-Net model

In line with the observed contrast properties, both intra-rater (ICC = 0.92, DSC = 0.87 ± 0.03) and inter-rater metrics (ICC = 0.83, DSC = 0.81 ± 0.03) indicated high reliability of manual CP segmentations on DIR sequences (Table 3).

**Table 3:** Reliability and performance metrics for CP segmentation on DIR in the internal test set (*n* = 20). ICCs are given with 95%-confidence intervals. Other values are expressed as mean ± standard deviation. Asymmetrically defined metrics have not been calculated for manual rater comparisons. Similarly, ICCs have not been determined for nnU-Net performance. CP, choroid plexus; DIR, double inversion recovery; DSC, Dice similarity coefficient; ICC, intraclass correlation coefficient; HD, Hausdorff distance; MSD, mean surface distance; RVD, relative volume difference.

|  | Intra-rater agreement | Inter-rater agreement | nnU-Net vs. manual |
| --- | --- | --- | --- |
| ICC | 0.92 (0.81–0.97) | 0.83 (0.63–0.93) | — |
| DSC | 0.87 ± 0.03 | 0.81 ± 0.03 | 0.82 ± 0.02 |
| Recall | — | — | 0.81 ± 0.04 |
| Precision | — | — | 0.83 ± 0.03 |
| MSD, mm | 0.21 ± 0.04 | 0.30 ± 0.06 | 0.27 ± 0.04 |
| HD, mm | 3.3 ± 1.3 | 4.2 ± 1.7 | 3.8 ± 1.6 |
| RVD, % | — | — | -2.6 ± 7.5 |
| Absolute RVD, % | 7.6 ± 6.3 | 12.4 ± 6.5 | 6.9 ± 3.6 |

Performance metrics of the nnU-Net segmentation model proved robust (DSC = 0.82 ± 0.02) and did not give reason for concern regarding systematic biases (recall = 0.81 ± 0.04, precision = 0.83 ± 0.03; Table 3). Distance-based and volume deviation metrics of the model were comparable to those observed for repeated segmentations by a single rater (MSD: nnU-Net = 0.27 ± 0.04 mm vs. intra-rater = 0.21 ± 0.04 mm, Hausdorff: nnU-Net = 3.8 ± 1.6 mm vs. intra-rater = 3.3 ± 1.3 mm, absolute RVD: nnU-Net = 6.9 ± 3.6% vs. intra-rater = 7.6 ± 6.3%). Furthermore, model performance (as measured by DSC) was not noticeably influenced by either scanner (*p* = 0.72), subject status (MS or HC, *p* = 0.77), or manual segmentation volume (*p* = 0.53). Finally, no proportional bias was detected, as the absolute volumetric difference between the nnU-Net and manual segmentations was not associated with the average CP volume (*p* = 0.84; Figure 3).

**Figure 3:**
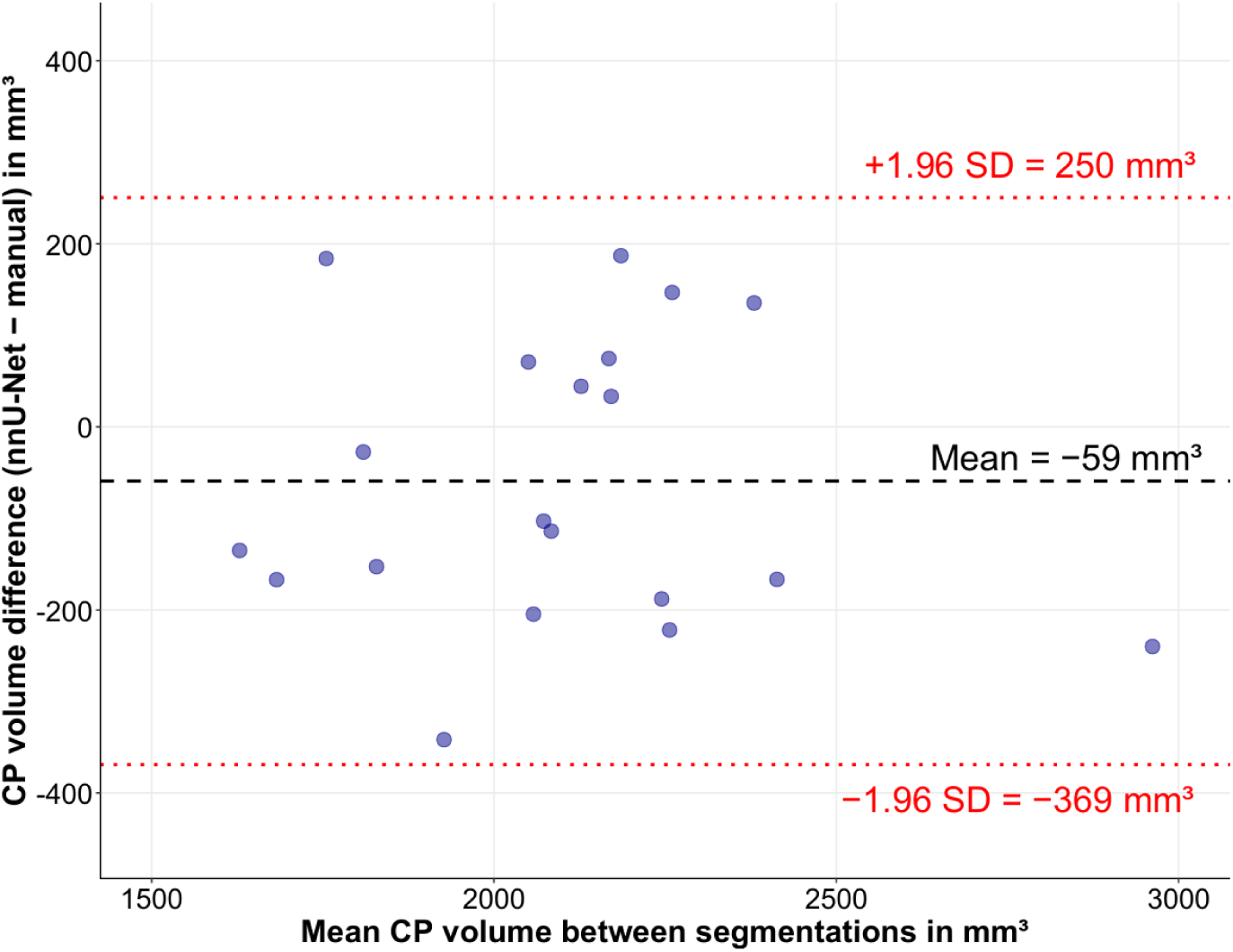
Bland-Altman plot for manual and nnU-Net CP volumes in the internal test set (*n* = 20). CP, choroid plexus; SD, standard deviation.

### Comparison with publicly available models

Mean CP volumes for manual segmentations (2133 ± 314 mm^3^) were comparable to those yielded by the nnU-Net model (2074 ± 307 mm^3^; *p* = 0.47). DIR-derived volumes for both manual and nnU-Net segmentations were significantly larger than T1w-derived ones (Li 2025: 1488 ± 411 mm^3^, Visani 2024: 1470 ± 573 mm^3^; all *p* < 0.01). Although manual DIR-based segmentation volumes were also significantly larger than FLAIR-based ones (Eisma 2024: 1826 ± 441 mm^3^; *p* = 0.02), this was only marginally true for nnU-Net volumes (*p* = 0.05). While the relative variance in CP volume was not significantly different between the manual, nnU-Net, Li 2025, or Eisma 2024 segmentations (all *p* > 0.05), the Visani 2024 model showed considerably higher variance than both DIR-based approaches (manual and nnU-Net) as well as the Eisma 2024 model (all *p* < 0.01). Overall, correlations of CP volumes between the different segmentations proved heterogeneous: Stronger correlations were observed between manual and nnU-Net volumes (*r* = 0.87) as well as between volumes yielded by the Li 2025 and Eisma 2024 models (*r* = 0.72; Figure 4A). Manual and nnU-Net CP volumes correlated moderately with both Li 2025 (*r* = 0.51 and *r* = 0.49, respectively) and Eisma 2024 volumes (*r* = 0.51 and *r* = 0.55, respectively). The model by Visani 2024 showed the weakest correlations with other segmentation approaches (*r* = 0.16 with manual, *r* = 0.19 with nnU-Net, *r* = 0.31 with Li 2025, and *r* = 0.07 with Eisma 2024).

**Figure 4:**
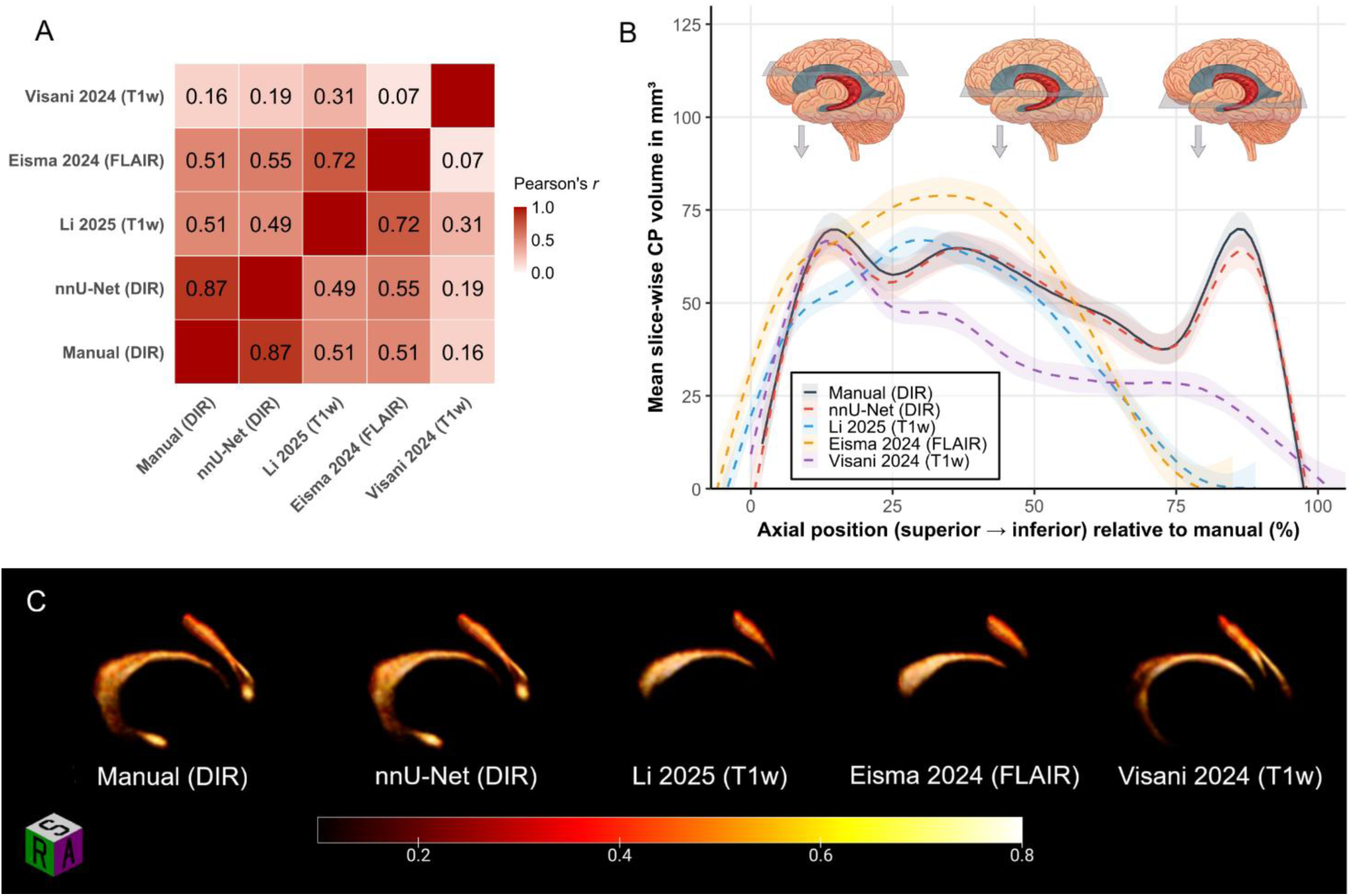
Manual segmentation of the CP vs. nnU-Net and publicly available models in the internal test set (*n* = 20). (A) Correlation matrix for CP volumes yielded by the different segmentation models. (B) Mean slice-wise CP volumes yielded by the different segmentation models transversing the brain axially from superior to inferior. The relative location of each slice is given as percentage relative to the manual segmentation. Shaded areas around the curves represent 95%-confidence intervals. (C) Heatmaps showing the frequency of a given voxel in MNI space being segmented by each of the models. A, anterior; CP, choroid plexus; DIR, double inversion recovery; FLAIR, fluid-attenuated inversion recovery; R, right; S, superior; T1w, T1-weighted.

Slice-wise analysis of axial CP volume distributions showed a volume spike toward the inferior/temporal part of the CP for the DIR-based segmentation approaches (manual and nnU-Net), which was not present for either the Li 2025 or Eisma 2024 models and only partly registered by the Visani 2024 model (Figure 4B). This graphical impression was confirmed by CDF analysis: The mean area under the CDF indicated a comparatively inferior-heavy (in relation to the z-axis) distribution for both DIR-based segmentations (manual: 53.9 ± 3.2%, nnU-Net: 53.9 ± 3.3%; *p* = 0.99), whereas the other models showed volume distributions that were significantly more concentrated toward superior slices (Visani 2024: 62.3 ± 5.4%, Li 2025: 68.0 ± 5.4%, Eisma 2024: 69.4 ± 4.8%; *p* < 0.001 against DIR-derived segmentations in each case). Heatmaps visualizing these local volume differences between segmentation models are shown in Figure 4C.

### External validation and generalizability

To evaluate the generalizability of our DIR-based nnU-Net model, its performance was assessed using an external dataset acquired on an MRI scanner from a different vendor (*n* = 10). While overlap and volume consistency metrics were worse than those seen in the internal test set, they nonetheless indicated overall generalizability of the nnU-Net model (DSC = 0.75 ± 0.02, absolute RVD = 15.2 ± 3.7%), albeit with a tendency toward undersegmentation (recall = 0.71 ± 0.05, precision = 0.80 ± 0.05, RVD = −11.1 ± 11.6%; Supplementary table 2). No significant proportional bias was detected (*p* = 0.11).

## Discussion

In this study, DIR was found to offer superior CP signal intensity and tissue contrast compared to conventional MRI sequences like T1w or FLAIR. Leveraging this advantage, a DIR-based nnU-Net segmentation model showed high internal accuracy and robust external generalizability. Moreover, it captured the CP in its full temporal/inferior extent, which we found to be largely missed by public T1w-or FLAIR-based models.

The current landscape of CP imaging studies commonly relies on T1w sequences to determine the structure’s volume. However, we found the T1w normalized signal intensity of the CP and the CP-to-GM contrast to be relatively poor, especially when measured against the respective DIR metrics. Our data suggest that this difference in MRI signal behavior might directly impact CP segmentation accuracy, as the internal performance of our DIR-based nnU-Net compares favorably to overlap and volume consistency metrics reported for previously published T1w-based models, that were also evaluated on internal datasets (average DSC: 0.82 vs. 0.74 [9, 14, 30, 33–35]; average absolute RVD: 6.9% vs. 12.5% [30, 33, 35]).

The lack of an established MRI standard for visualizing the CP makes it difficult to evaluate the anatomical fidelity of different segmentation approaches. However, observations from pathology and surgery describe the CP as essentially C-shaped, extending from the body of the lateral ventricle (near the foramen of Monro) into its temporal horn along what is called the choroidal fissure [36, 37]. This stands in contrast to a number of recent publications, where 3D models reconstructed from CP segmentations tend to miss the lower part of the C-structure [13–16]. Consistent with this, three public T1w- and FLAIR-based segmentation models applied to our dataset yielded CP volumes that were 24% smaller on average than those derived from DIR-based approaches. Crucially, these public models only partly captured – or completely missed – voxels within the temporal horn of the lateral ventricle. While the impact of this incomplete coverage on the utility of CP volume as a biomarker remains unclear, it introduces a significant risk of volumetric bias: Enlarged lateral ventricles improve the visibility of the temporal CP on T1w and FLAIR scans, which may paradoxically result in a disproportionate reduction of under-segmentation errors in those specific patients. Of note, many T1w-based studies have noticed substantial positive correlations (*r* > 0.5) between CP and lateral ventricle volumes [12, 15, 38–42]. Although this association is pathophysiologically plausible in principle, it may be significantly confounded by the interaction between altered ventricular geometry and sequence-specific contrast limitations (reduced partial volume effects in enlarged ventricles artificially inflating the relative CP volume).

A primary barrier to the broader clinical translation of DIR-based CP volumetry is sequence availability; while DIR is increasingly standard in MS protocols, it is rarely acquired in routine evaluations for other CNS disorders. To bridge this gap, synthetic imaging offers a promising solution. Generated from standard T1w and T2-weighted scans using deep learning methods, synthetic DIR has primarily been evaluated for MS lesion detection. In that context, it significantly outperformed conventional MRI sequences [43, 44] and was generally comparable to physically acquired DIR sequences in both single-center [45] and multi-center settings [46]. Whether synthetic DIR can serve as a viable substitute in CP segmentation remains to be investigated.

This study has some limitations: The datasets used for model training and testing were restricted to HC and people with MS. This is essentially due to the fact that DIR sequences are not commonly available for other CNS disorders like dementia, stroke, or schizophrenia, where CP volume has also been discussed as a potential imaging biomarker. Nonetheless, including additional diagnoses might have improved the generalizability of our segmentation model. Similarly, the training data was confined to three different 3 Tesla scanners by the same manufacturer. While there are advantages to this approach – allowing the network to extract core anatomical features of the CP without being confounded by hardware-related variance – a more heterogeneous training pool might have improved the model’s robustness to variations in image contrast, field strength, and scanner-specific artifacts.

Indeed, this domain-shift vulnerability is reflected in our external validation, where the model demonstrated a drop in spatial overlap (DSC: 0.82 vs. 0.75) and a notable decrease in volume consistency (absolute RVD: 6.9% vs. 15.2%). This performance degradation on external data may impede clinical biomarker translation and require dataset-specific fine-tuning prior to application – an option we have made available as part of our public segmentation model. Finally, we found the contrast between the CP and adjacent lesional tissue on DIR to be comparatively low. This overlapping hyperintense signal profile may render DIR-based CP segmentation challenging in certain scenarios, particularly in neuroinflammatory disorders with extensive periventricular demyelination or in older individuals with pronounced age-related WM hyperintensities. While we addressed this vulnerability by manually correcting misclassified edge cases within an active-learning framework, we cannot entirely rule out the risk of localized segmentation errors or partial volume artifacts in patients with high lesion loads.

In conclusion, DIR offers distinct advantages for CP segmentation over conventional MRI sequences, primarily with respect to segmentation consistency and complete coverage of CP anatomy. This is reflected in superior performance metrics of an nnU-Net model based on DIR-derived CP masks, which we have made publicly available. Our findings may be an important step toward standardizing CP volumetry to facilitate the evaluation of its biomarker potential in future studies on CNS disorders.

## List of abbreviations

ANOVA: Analysis of variance
CDF: Cumulative distribution function
CE: Contrast-enhanced
CNS: Central nervous system
CoV: Coefficient of variation
CP: Choroid plexus
CSF: Cerebrospinal fluid
DIR: Double inversion recovery
DSC: Dice similarity coefficient
FLAIR: Fluid-attenuated inversion recovery
GM: Gray matter
HC: Healthy controls
HD: Hausdorff distance
HSD: Honestly Significant Difference (Tukey’s test)
ICC: Intraclass correlation coefficient
MRI: Magnetic resonance imaging
MS: Multiple sclerosis
MSD: Mean surface distance
RVD: Relative volume difference
SD: Standard deviation
T1w: T1-weighted
TUM: Technical University of Munich
WM: White matter

## Data Availability

The anonymized data that supported the findings of this study are available upon reasonable request. The segmentation model developed in the study has been made publicly available at https://github.com/mlauerer/choroid-plexus-segmentation-dir.

## Acknowledgments

We thank the study participants for their invaluable contributions and the clinic support staff for their technical and administrative assistance. We also express our gratitude to Achim Berthele for curating the TUM-MS cohort.

## Availability of data and materials

The anonymized data that supported the findings of this study are available from the corresponding author upon reasonable request. The model weights and instructions for deploying the nnU-Net are publicly available at https://github.com/mlauerer/choroid-plexus-segmentation-dir.

## Authors’ contributions

M.L., J.M., C.B., T.W., and M.M. contributed to the conception and design of the study, as well as data analysis. P.B., E.A.H., J.S.K., and B.H. participated in the acquisition of data. M.L. and M.M. drafted the text and figures.

