## Supplementary Material for "Improved visualization and segmentation of the choroid plexus using double inversion recovery MRI"

**Supplementary table 1:** Detailed MRI sequence parameters across the four study scanners

**Supplementary table 2:** Performance metrics for CP segmentation on DIR in the external test set (*n* = 10)

**Supplementary figure 1:** Representative segmentation errors after first round of nnU-Net training

**Supplementary figure 2:** Example slices for CP contrast on different MRI sequences

### Supplementary table 1: Detailed MRI sequence parameters across the four study scanners

| | Achieva (Philips) | Achieva dStream (Philips) | Ingenia (Philips) | Skyra (Siemens) |
|---|---|---|---|---|
| Field strength | 3 Tesla | 3 Tesla | 3 Tesla | 3 Tesla |
| **3D DIR** | | | | |
| TR (ms) | 5,500 | 5,500 | 5,500 | 5,000 |
| TE (ms) | shortest | shortest | shortest | shortest |
| TI (ms) | 2,550 | 2,550 | 2,550 | 1,800 |
| FA (°) | 90 | 90 | 90 | 120 |
| Voxel size ($mm^3$) | 1.0 × 1.0 × 1.3 | 0.75 × 0.75 × 0.75 | 0.75 × 0.75 × 0.75 | 0.45 × 0.45 × 0.90 |
| **3D T1w (MPRAGE)** | | | | |
| TR (ms) | 9 | 9 | 9 | — |
| TE (ms) | 4 | 4 | 4 | — |
| TI (ms) | 1,000 | 1,000 | 1,000 | — |
| FA (°) | 8 | 8 | 8 | — |
| Voxel size ($mm^3$) | 1.0 × 1.0 × 1.0 | 0.75 × 0.75 × 0.75 | 0.75 × 0.75 × 0.75 | — |
| **3D FLAIR** | | | | |
| TR (ms) | 10,000 | 4,800 | 4,800 | — |
| TE (ms) | shortest | shortest | shortest | — |
| TI (ms) | 2,750 | 1,650 | 1,650 | — |
| FA (°) | 90 | 90 | 90 | — |
| Voxel size ($mm^3$) | 0.9 × 0.9 × 1.5 | 0.75 × 0.75 × 0.75 | 0.75 × 0.75 × 0.75 | — |

No sequences other than DIR were analyzed in the external Siemens dataset. DIR, double inversion recovery; FA, flip angle; MRI, magnetic resonance imaging; TE, echo time; TI, inversion time; TR, repetition time.

### Supplementary table 2: Performance metrics for CP segmentation on DIR in the external test set ($n$ = 10)

| | nnU-Net vs. manual |
|---|---|
| Dice similarity coefficient | 0.75 ± 0.02 |
| Recall | 0.71 ± 0.05 |
| Precision | 0.80 ± 0.05 |
| Mean surface distance, mm | 0.27 ± 0.03 |
| Hausdorff distance, mm | 5.3 ± 1.8 |
| Relative volume difference, % | −11.1 ± 11.6 |
| Absolute relative volume difference, % | 15.2 ± 3.7 |

Values are expressed as mean ± standard deviation.

### Supplementary figure 1: Representative segmentation errors after first round of nnU-Net training

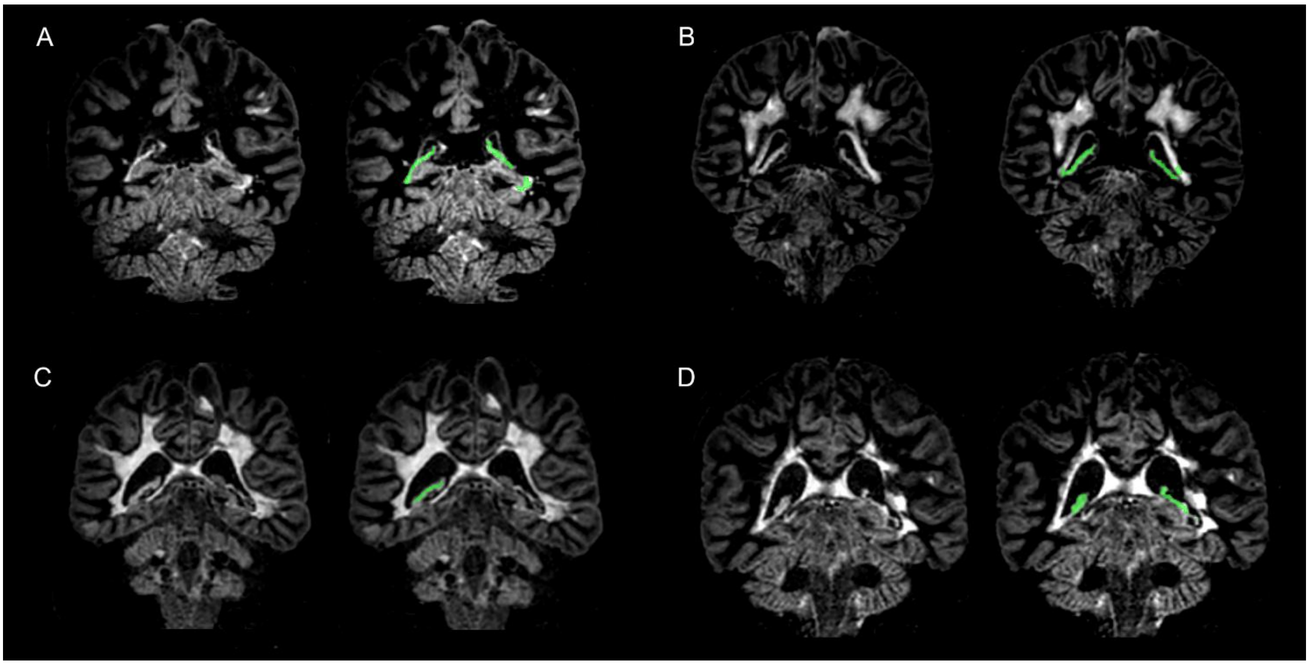


Coronal DIR slices of individuals with MS, nnU-Net segmentations are shown in green. (A) Misclassification of a hyperintense periventricular white matter lesion as CP tissue, immediately adjacent to the temporal horn of the left lateral ventricle. (B) Erroneous segmentation of periventricular white matter lesion tissue at the left atrium of the lateral ventricle. (C) Substantial undersegmentation of CP tissue in a brain with global atrophy and extensive periventricular white matter lesions. (D) Missed segmentation of inferior CP tissue in a brain with enlarged ventricles and considerable periventricular lesion tissue. CP, choroid plexus; DIR, double inversion recovery; MS, multiple sclerosis.

**Supplementary figure 2: Example slices for CP contrast on different MRI sequences**

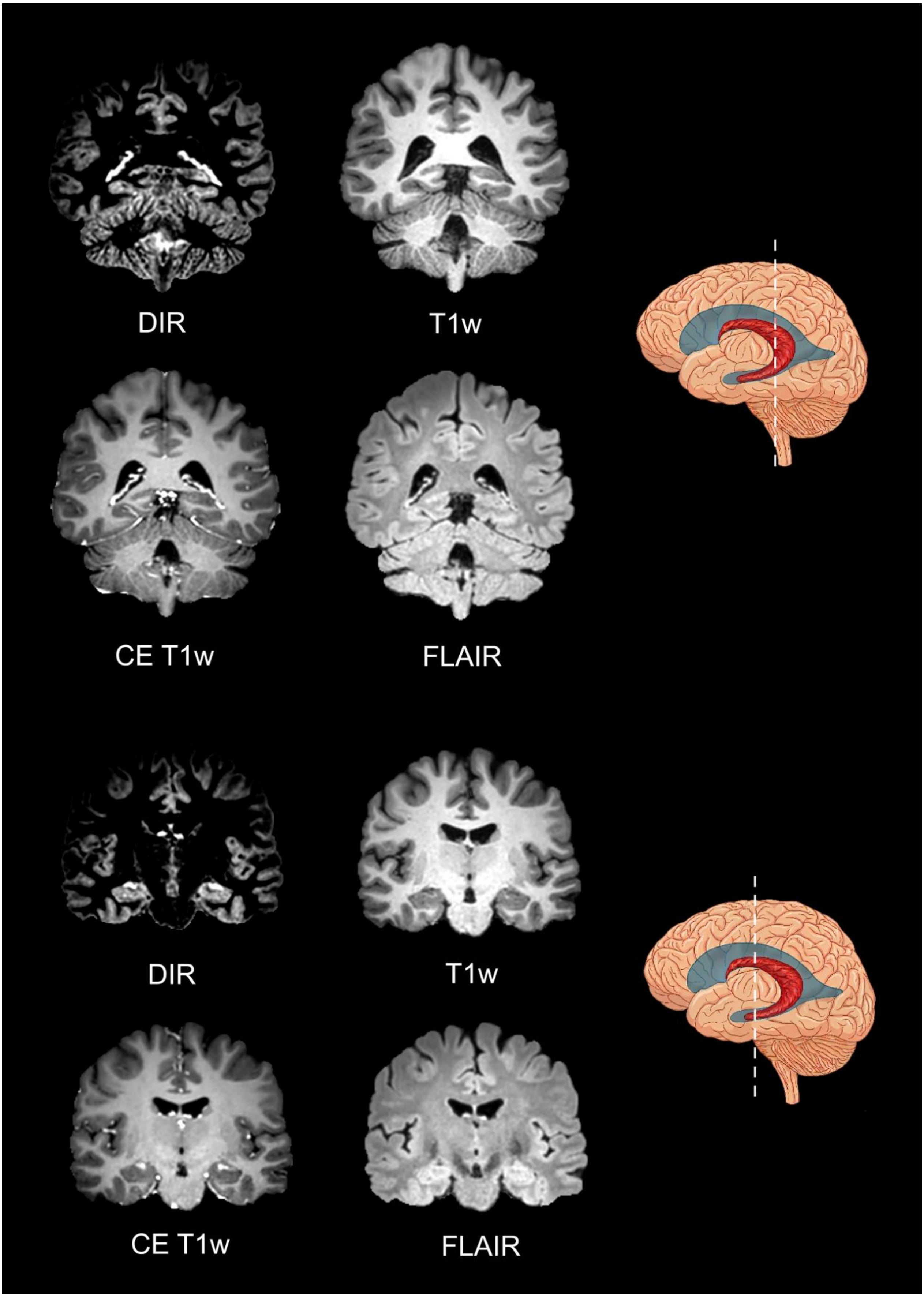


Coronal brain slices visualizing sequence-specific CP contrast at two different anatomical locations. CE, contrast-enhanced; CP, choroid plexus; DIR, double inversion recovery; FLAIR, fluid-attenuated inversion recovery; T1w, T1-weighted.
